# Clinical value of cortical bursting in preterm infants with intraventricular haemorrhage

**DOI:** 10.1101/2023.04.25.23289131

**Authors:** Tuomas Koskela, Judith Meek, Angela Huertas-Ceballos, Giles S Kendall, Kimberley Whitehead

## Abstract

**Objective:** In healthy preterm infants, cortical burst rate and temporal dynamics predict important measures such as brain growth. We hypothesised that in preterm infants with germinal matrix-intraventricular haemorrhage (GM-IVH), cortical bursting could provide prognostic information. We determined how cortical bursting was influenced by the injury, and whether this was related to developmental outcome.

**Methods:** We identified 47 EEGs from 33 infants with GM-IVH ≥grade II (median gestational age: 25 weeks), acquired between 24-40 weeks corrected gestational age as part of routine clinical care. In a subset of 33 EEGs from 25 infants with asymmetric injury, we used the least-affected hemisphere as an internal control. We tested whether cortical burst rate predicted death or severe motor impairment (median 2 years follow-up; range 1-2 years corrected).

**Results:** GM-IVH depressed central cortical burst rate. Bursts over the worst-affected hemisphere were less likely to immediately follow (within 1 second) bursts over the least-affected hemisphere than vice versa. Lower burst rate was modestly associated with death or severe motor impairment (specificity 93%, sensitivity 37%).

**Conclusions:** EEG can quantitatively index the functional injury after GM-IVH.

**Significance:** Higher cortical burst rate is reassuring for a positive motor outcome over the first 2 years.

**Highlights:**

- GM-IVH depresses cortical burst rate
- GM-IVH unbalances inter-hemispheric burst dynamics
- Higher burst rate following GM-IVH is associated with positive motor outcome at median 2 years

## 1. Introduction

In preterm infants, the largest burden of acquired brain injury is intraventricular haemorrhage arising from the germinal matrix (GM-IVH) (Ancel et al., 2015; Gale et al., 2018; de Vries, 2018). This injury can occur spontaneously, or be triggered or exacerbated by acute illness such as sepsis or metabolic acidosis (Horst et al., 2010; Linder et al., 2003; Thorp et al., 2001). The injury is graded on a four-point ascending scale of severity, depending on the worst of serial cranial ultrasound scans. GM-IVH of grade II or higher is associated with worse outcomes relative to gestational age-matched controls, with ventricular dilatation and intraparenchymal involvement conferring additional risk of disability or death (Bolisetty et al., 2014; Cizmeci et al., 2020; Hollebrandse et al., 2021; Mukerji et al., 2015; Patra et al., 2006). However, the grade of structural injury only coarsely predicts outcome, e.g. 20% of infants with intraparenchymal damage have entirely normal outcome (Marret et al., 2013). Complementary prognostic information would help to direct early therapeutic intervention.

At University College London Hospitals (UCLH), infants with GM-IVH are monitored with electroencephalography (EEG) when seizures are suspected, which are known to be associated with the injury (Hellström-Westas et al., 2001, 1991). Additionally, it is possible that the background EEG could contribute to neurological assessment and prognostication, as is customary following brain injury in full-term neonates (Ouwehand et al., 2020). The dominant feature of the neonatal EEG is cortical bursting activity. Cortical bursts reflect excitatory input to pyramidal neurons in animal models (Golbs et al., 2011; Hanganu et al., 2007; Minlebaev et al., 2009). Burst rate and dynamics in healthy preterm infants predict subsequent brain growth and microstructure, as well as mental development (Benders et al., 2015; De Wel et al., 2021; Guzzetta et al., 2009; Iyer et al., 2015; Tataranno et al., 2018). Cortical bursts can be attenuated by GM-IVH (Aso et al., 1993, 1989; Chalak et al., 2011; Connell et al., 1988; Greisen et al., 1987; Olischar et al., 2007; Ranasinghe et al., 2015). Thus, we hypothesised that their rate of occurrence and temporal dynamics could index the functional injury and potentially be prognostic.

## 2. Methods

### 2.1 Infants

This project was defined as a retrospective service evaluation by the UCLH Research and Development Directorate and therefore individual consent from parents was not required. All clinical data review was conducted by a UCLH-affiliated, state-registered Clinical Neurophysiologist (KW). We identified infants born between 2007 and 2022 who underwent EEG monitoring during the neonatal period (defined here as ≤40 weeks corrected gestational age (CGA)) which was available for review. Selection criteria comprised gestational age <35 weeks (Ancel et al., 2015; Aso et al., 1993; Connell et al., 1988; Radvanyi-Bouvet et al., 1987) and evidence of ≥grade II GM-IVH on routine cranial imaging. Exclusion criteria included evidence of intrapartum hypoxic-ischemic insult, or acute severe metabolic disturbance at the time of EEG. This resulted in a total sample of 34 infants with median gestational age 25+5 weeks+days.

### 2.2 EEG monitoring for suspected seizures

A minimum of 4 Ag/AgCl recording electrodes were positioned at bilateral central and frontal sites (C4, C3, F4, F3), according to the international 10/20 electrode placement system. Eight/34 infants had more than one EEG. This resulted in a total of 52 recordings (Table 1), which were all reviewed for electrographic seizures (Pressler et al., 2021).

**Table 1:** Infant demographics.

|  |  |
| --- | --- |
| Total no. of infants with GM-IVH | 34 |
| Grade II | 7/34 |
| Ventricular dilatation<sup>a</sup> | 5/34 |
| With intraparenchymal lesion(s) | 22/34 |
| *Total asymmetric injury* | *26/34 (5 grade II, 2 ventricular dilatation, 19 with intraparenchymal lesion(s))* |
| Sex (female: male) | 12:22 |
| Median (range) birth weight (grams) | 821 (533 - 1999) |
| Median (range) gestational age (weeks+days) | 25+5 (23+4 - 34+2) |
| Median (range) Apgar score at 1 minute | 4 (1 - 9) |
| Median (range) Apgar score at 5 minutes | 8 (2 - 10) |
| No. of EEGs | 52 EEGs from 34 infants |
| Median (range) corrected gestational age (weeks+days) at discharge home from hospital (in survivors) | 42+6 (36+5 - 56+3) |
| No. of EEGs suitable for analysis of cortical bursting<sup>b</sup> | 47 EEGs from 33 infants |
| Median (range) postnatal age (days) | 20 (2 - 104) |
| Median (range) corrected gestational age (weeks+days)<sup>c</sup> | 30+1 (24+0 - 40+1) |
| Morphine exposure | 27/47 |
| Anti-seizure drug exposure<sup>d</sup> | 16/47 |

<sup>a</sup> Defined here as dilatation $\geq 97^{\text{th}}$ centile and/or anterior horn width $> 6\text{mm}$ (Kidokoro et al., 2014; Leijser et al., 2018; Vries et al., 2002).
<sup>b</sup> The infant without any EEG suitable for bursting analysis died on postnatal day 1; they had asymmetric GM-IVH with intraparenchymal lesion. In cases of $> 1$ EEG being analysed from the same infant, the mean inter-recording interval was 10 days, which does not underestimate the variance of EEG-level analyses (see the supplemental information in (Fabrizi et al., 2011)).
<sup>c</sup> At which analysed EEG ended. Corrected gestational age = Gestational age + postnatal age.
<sup>d</sup> 15 Phenobarbital, 3 Benzodiazepine, 2 Phenytoin, 1 Levetiracetam, 1 Paraldehyde. (Total adds up to $> 16$ because 6 EEGs were acquired during exposure to 2 anti-seizure drugs).

### 2.3 EEG analysis of cortical bursting

#### 2.3.1 Inclusion criteria for EEG analysis of cortical bursting

For the analysis of cortical bursting, we excluded 5 recordings with >20% seizure burden (Payne et al., 2014) and/or during treatment with 3 anti-seizure drugs, which precluded appraisal of background EEG features (Arkilo et al., 2013; Osredkar et al., 2005; Ranasinghe et al., 2015). In the remaining 47 recordings, we selected segments with no or fewer seizures, including up to 2 days of data per recording: 41/47 segments included no seizures, and the remaining 6 segments had ≤0.5% seizure burden.

#### 2.3.2 Burst occurrence rate

Cortical bursts which comprise fast oscillations (8-30Hz) nested into slower rhythms are the dominant background pattern of the neonatal EEG (Whitehead et al., 2016). To identify these bursts, we first removed artefactual sections by visual inspection using ‘pop_eegplot’ in EEGLAB v.14 (Delorme and Makeig, 2004). We then calculated root-mean-square (RMS) amplitude values between 8-30Hz, using sliding 400-ms intervals (Antony et al., 2018; Hartley et al., 2012; Ranasinghe et al., 2015). We identified segments for each channel that were consecutively above a set threshold (1.5 times the standard deviation of its RMS signal over the whole recording (Antony et al., 2018; Vanhatalo et al., 2005)) for ≥0.5 seconds (Conde et al., 2005; Omidvarnia et al., 2014), using ‘detectevent’ in EEGLAB (for illustration, see (Koskela et al., 2021b)). Please see Supplementary Information for further details about data pre-processing.

#### 2.3.3 Burst temporal dynamics

The relative timing of cortical bursts overlying different brain regions offers insight into functional connections (Leikos et al., 2020; Tokariev et al., 2012). To examine whether GM-IVH influenced the temporal relationship between burst onsets at recording channels overlying different regions, we represented their latencies with a gaussian window of 3 standard deviations around each value, using ‘gauss’ in EEGLAB. We then calculated cross-correlations for 6 positive and 6 negative lag values between -1500 and +1500 ms, normalised to the autocorrelation between identical burst latencies (i.e. correlation of 1.00 at lag 0 ms) (Hartley et al., 2012; Koskela et al., 2021a; Leroy-Terquem et al., 2017).

#### 2.3.4 Burst magnitude

The magnitude of bursts can be indexed by their power (μV^2^). To characterise power changes of detected bursts relative to baseline, we convoluted the EEG signal with a Morlet wavelet between 0.1–45Hz using an increasing range of cycles (3–270), employing ‘newtimef’ in EEGLAB. For bursts detected at each channel, we extracted the 8-30Hz power at that channel over the course of the burst, and then normalised this value by dividing by its duration in seconds (Koskela et al., 2021a).

#### 2.3.5 Cortical bursting and motor outcome

Positive neurodevelopmental outcome was specified as survival without severe impairment (defined here as Bayley Scales of Infant Development 3^rd^ edition motor composite score ≤70/100, or unable to be assessed using Bayley Scales because of severe global delay and cerebral visual impairment; median 2 years follow-up, range 1-2 years corrected).

### 2.4 Statistical analysis

To assess differences between matched intra-subject variables or unpaired variables we used paired and unpaired t tests respectively. To test for associations between two continuous variables we used Pearson correlations.

To investigate multiple factors potentially underlying variance in cortical bursting, we conducted a hierarchical linear regression in which CGA was entered as the first explanatory variable given its known large effect (Whitehead et al., 2016). After that, we examined whether adding further EEG-level variables of morphine or anti-seizure drug(s) exposure, or electrographic seizure(s) during the analysed segment (all yes/no) improved model fit (Bell et al., 1993; Ranasinghe et al., 2015; Tataranno et al., 2020). Finally, we tested whether adding the infant-level variable of intraparenchymal lesion(s) optimised model fit.

To examine whether cortical bursting predicted survival without impairment, we conducted a receiver operating characteristic (ROC) analysis and i) calculated the area under the curve (AUC) which is a combined measure of sensitivity and specificity: an AUC of 0.5 indicates prediction no better than chance and serves as the null hypothesis, while 1.0 would reflect a perfect predictor, and then ii) examined ROC curve coordinates to identify cut-off thresholds which were optimally predictive. Statistical analysis was performed using IBM SPSS v. 26 and significance was set at p < .05.

## 3. Results

### 3.1 Seizures

At the EEG-level, 17/52 (33%) EEGs included electrographic seizures. At the infant-level, 15/34 (44%) infants had electrographic seizures recorded during at least one EEG. These seizures were recorded between postnatal days 0-104 and 25-40 weeks CGA, in line with previous reports that seizures can occur many weeks after the initial injury (Pisani et al., 2018, 2008; Scher et al., 1993). Please see Supplementary Fig. 1 for a seizure example.

### 3.2 Characterisation of cortical bursts

In 33/34 infants, at least one EEG was suitable for bursting analysis (see Table 1 for demographic and medication information). The 47 analysed EEGs were of median duration 8 hours (minimum 0.5 hours in 45/47 recordings). Cortical bursts had mean duration 1.6 seconds. These bursts comprised an increase in power which peaked between 8-30Hz as expected, coupled to a less pronounced but longer-duration increase in slower frequencies (Fig. 1). For bursts identified at each channel, the largest changes in 8-30Hz power were at that channel as anticipated, although bursts involved other channels also, especially for slower frequencies (Fig. 1).

**Fig. 1:**
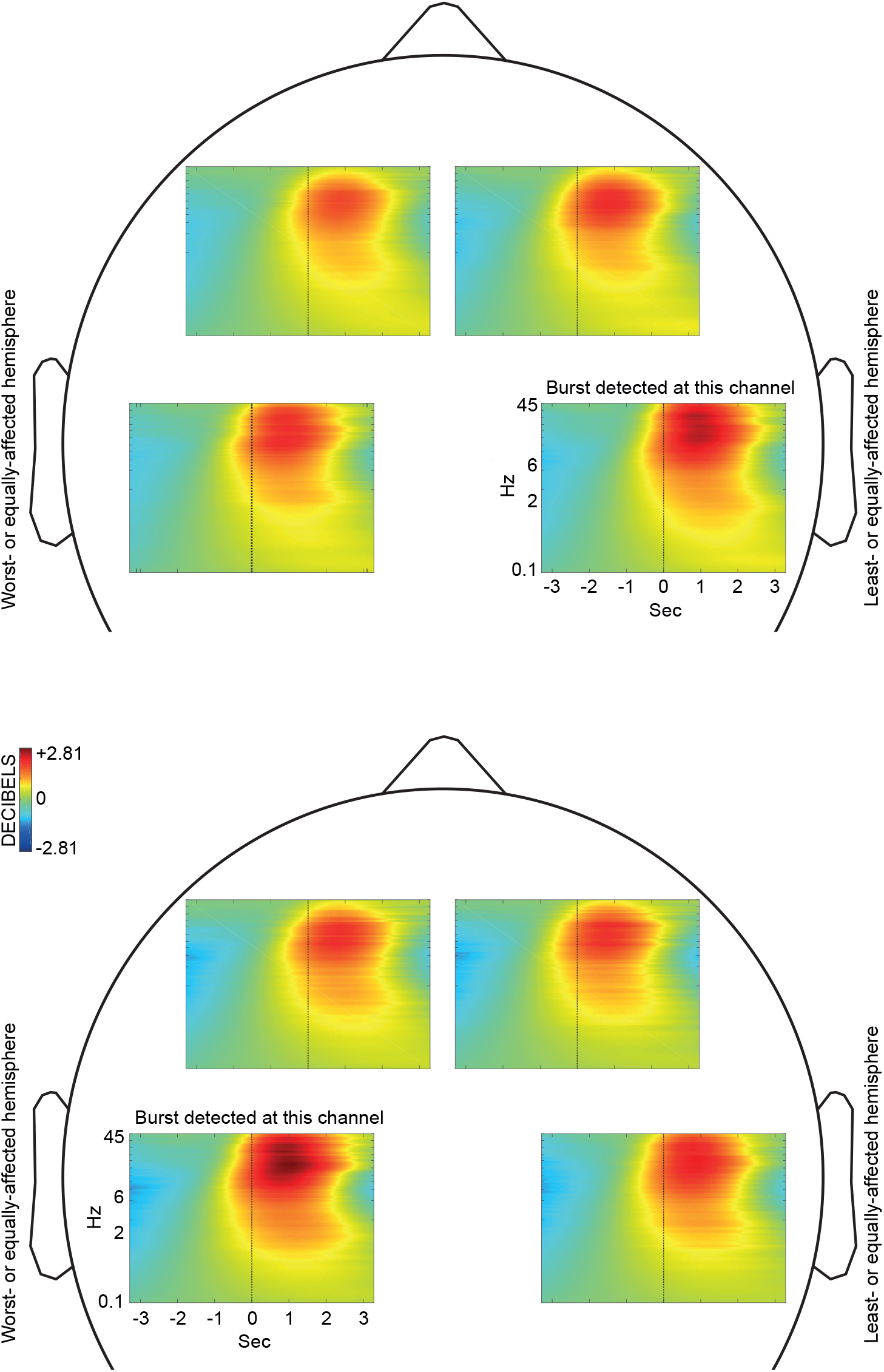
Grand average time-frequency changes associated with cortical bursts. Bursts identified at the central channel overlying the least-affected brain hemisphere (or right hemisphere in the case of symmetric injury) (upper panel) and worst-affected brain hemisphere (or left hemisphere in the case of symmetric injury) (lower panel). Power changes between 0.1-45Hz (logarithmic scale) are shown in decibels, relative to the mean power preceding burst onset (black vertical line), where increased power is red and decreased power is blue.

### 3.3 Cortical burst rate was depressed by GM-IVH

To examine whether injury altered cortical burst rate, we first took advantage of a subgroup of 33 EEGs from 25 infants with asymmetric injury, for whom we could use the least-affected hemisphere as an internal control. There were fewer central (but not frontal) bursts per minute over the worst-affected hemisphere (central: mean 7.9 vs. 9.3, [95% CI of difference -2.10 -0.62], p = .001, Fig. 2; frontal: p = .648). This inter-hemispheric difference in central burst rate did not significantly narrow with postnatal age (p = .415) or CGA (p = .169, Fig. 2). In comparison and as expected, there was no inter-hemispheric difference in central burst rate in symmetric injury (p = .204, Fig. 2), when inter-hemispheric bursting ratio was more equal than in asymmetric injury (mean ratio 1.04 vs. 0.85, [95% CI of ratio difference 0.09 0.28], p < .001).

**Fig. 2:**
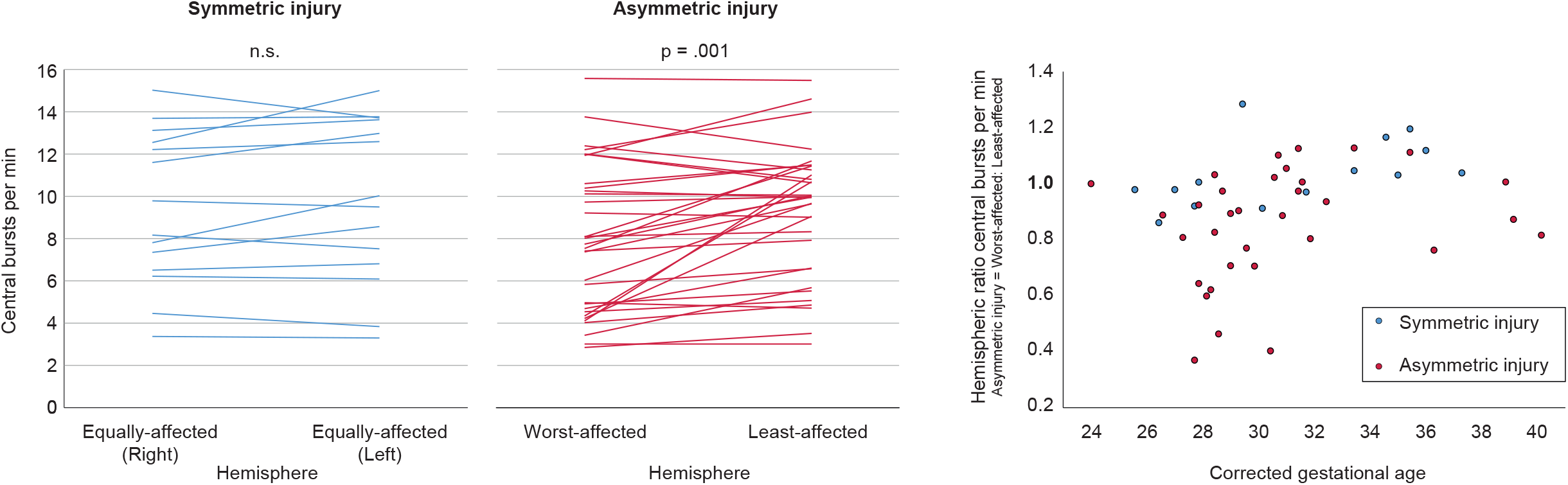
Cortical burst rate was depressed by GM-IVH. Left: Central burst rate over the two hemispheres when injury is symmetric (n = 14 EEGs) or asymmetric (n = 33 EEGs). Each EEG is represented by one line. Right: Scatter plot of inter-hemispheric burst rate ratio against corrected gestational age at EEG. For infants with symmetric injury, the hemispheric ratio is left: right hemisphere. Each EEG is represented by one dot.

Central bursts over the worst-affected hemisphere were less likely to follow bursts over the least-affected hemisphere than vice versa (negative lags had lower cross-correlations than their paired positive lag between 250-1000 ms (e.g. -500 vs. +500 ms) (p ≤ .014, Fig. 3). In comparison and as expected, in symmetric injury inter-hemispheric central burst onsets were balanced (no significant differences between the cross-correlations of paired lags: p ≥ .374, Fig. 3).

**Fig. 3:**
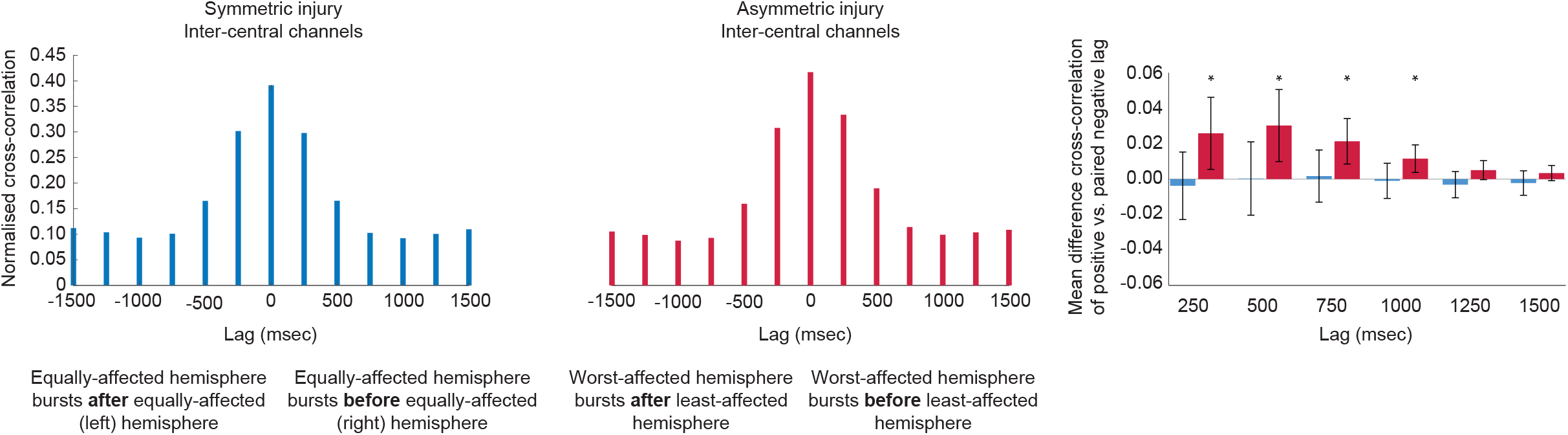
Inter-hemispheric cortical burst dynamics were altered by asymmetric GM-IVH. The relative timing of central burst occurrence over the two hemispheres when injury is symmetric (n = 14 EEGs) or asymmetric (n = 33 EEGs). Central bursts over the worst-affected hemisphere were less likely to immediately follow bursts over the least-affected hemisphere than vice versa. * = p < .05. 95% confidence intervals are denoted by error bars.

In asymmetric injury, greater depression of central burst rate predicted higher mean central burst power at the worst-affected hemisphere (inter-hemispheric ratio of central burst rate vs. power: r = -.608, p < .001), but not the least-affected hemisphere (p = .350). Pooling EEGs from all infants together, with either asymmetric or symmetric injury, also showed an association between lower burst rate and higher mean burst power at that same region (r = -.390 to -.569, p ≤ .007).

### 3.4 Lower cortical burst rate was associated with adverse outcome

Central burst rate over the worst- or equally-affected hemisphere increased with CGA, and was slightly reduced by morphine exposure (but not by anti-seizure drug exposure, or proximal electrographic seizures) (Table 2). We reasoned that lower burst rate than expected after accounting for these factors could reflect worse functional injury, and predict adverse outcome. To test this, we calculated standardised residuals (z-scores) after fitting the CGA + morphine model: a z-score above 0 indicates that burst rate was higher than predicted by the model, a z-score below 0 indicates that burst rate was lower than predicted by the model.

**Table 2:** Hierarchical linear models of variables influencing central burst rate over the worst- or equally-affected hemisphere.

|  | B [95% CI] | s.e. | *p* | R<sup>2</sup> |
| --- | --- | --- | --- | --- |
| *Model 1: CGA* |  |  |  |  |
| CGA | .637 [.412 .861] | .111 | <.001 | .421 |
| *Model 2: CGA + morphine<sup>a</sup>* |  |  |  |  |
| CGA | .596 [.382 .810] | .106 | <.001 | .495 |
| Morphine | -2.046 [-.427 -3.664] | .803 | .014 |  |
| *Model 3: CGA + morphine + IPL* |  |  |  |  |
| CGA | .639 [.433 .845] | .102 | <.001 | .557 |
| Morphine | -2.114 [-.578 -3.650] | .762 | .008 |  |
| IPL | -2.007 [-.351 -3.664] | .822 | .019 |  |

<sup>a</sup> Anti-seizure drug exposure and Electrographic seizures were both excluded from the model: *p* = .168 and .301 respectively.
IPL = Intraparenchymal lesion(s)

Of 33 infants with bursting analysed, 27 infants - who had 41 EEGs in total - had outcome information available (six infants died after redirection of care, and outcome was available for 21/27 surviving infants). Lower central burst rate was modestly associated with adverse outcome (AUC .648, specificity and sensitivity 93% and 37% respectively using an optimal cut-off threshold of -0.77; Fig. 4a).

**Fig. 4:**
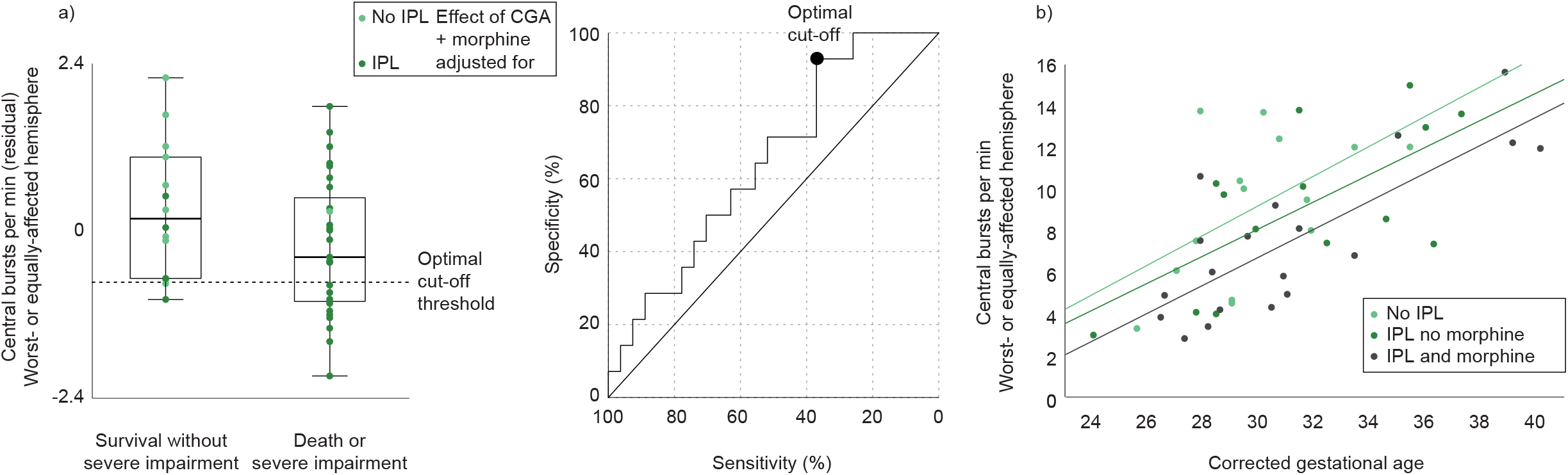
Burst occurrence rate and incidence of death or severe motor impairment. a) Distribution of standardised residual of burst occurrence rate after corrected gestational age and morphine exposure controlled for in infants who did or did not survive without severe motor impairment. Each EEG is represented by a dot (n = 41). The optimal cut-off threshold to predict death or severe impairment is represented by a black dashed line, and the adjacent ROC curve illustrates how this threshold was derived. Data are colour-coded by whether the infant had intraparenchymal lesion(s) (IPL), to demonstrate the degree to which this factor co-varied with burst rate and outcome. Right: For illustrative purposes, the cross-sectional developmental trajectory of burst rate is fitted separately for three subgroups to demonstrate how IPL and morphine exposure reduced burst rate.

After adding presence (yes/no) of intraparenchymal lesion(s) (IPL) to the CGA + morphine model explaining burst rate, model fit was improved because IPL attenuated burst rate (Table 2; visualised in Fig. 4b), and the residuals no longer predicted outcome (AUC .540). This suggests that the association between burst rate and outcome is partially mediated by whether GM-IVH is associated with IPL. However, Fig. 4a shows that burst rate can also provide unique information: in three instances of IPL but relatively high burst rate, the cut-off threshold correctly predicted that the infant did not suffer an adverse outcome.

## 4. Discussion

GM-IVH depresses burst rate over the sensitive period equivalent to the third trimester of gestation when cortical bursting refines neural circuits in animal models (Lebedeva et al., 2017; Molnár et al., 2020; Ranasinghe et al., 2015; Tolner et al., 2012). This is also when activity-dependent emergence of bilateral cortical networks occurs, which could be disrupted by skewed inter-hemispheric cortical burst dynamics when one hemisphere is injured relative to the other (Allievi et al., 2016; Marcano-Reik et al., 2010; Tokariev et al., 2012; Whitehead et al., 2022, 2019). These abnormalities of burst initiation (rate) and propagation (inter-hemispheric dynamics) are likely to reflect the grey matter and white matter damage associated with GM-IVH grade II or higher (Omidvarnia et al., 2015; Tortora et al., 2018; Vasileiadis et al., 2004).

Burst rate was modestly associated with outcome, which to our knowledge is the only recent report that the background EEG is prognostic in infants with GM-IVH, since two much earlier papers when neonatal intensive care was very different (Connell et al., 1988; Hellström-Westas et al., 2001). Therefore, EEG could be used to track prognostic information over time. This has the potential to provide real time monitoring of the effect of clinical interventions delivered on the neonatal unit after the injury, e.g. to support sleep cycling as it emerges from approximately 28-31 weeks CGA (Supplementary Fig. 2) (Georgoulas et al., 2021; van den Hoogen et al., 2017).

Our results indicate that the background EEG could contribute to neurological assessment and prognostication after brain injury in preterm infants, as is customary in full-term infants. Indeed, in both cohorts injury is associated with sparser, higher power cortical bursts, suggesting some similarity in how the insult impacts brain function (Koskela et al., 2021b; Lamblin et al., 2013; Whitehead et al., 2020).

This work has some limitations. The sample was varied, but this is a true reflection of our clinical population and much of the inter-subject heterogeneity was controlled for by use of intra-subject analyses. We successfully used this intra-subject approach to show that burst rate was depressed over the worst-vs. least-affected hemisphere, but a cleaner control would have been a matched group with no GM-IVH. Furthermore, the sample underwent EEG recordings because seizures were suspected; proximal seizures could contribute to the depression of inter-ictal cortical bursting and therefore the sample may not be representative of the total population of infants with GM-IVH. In the future, a multi-centre study could be conducted to model predictors of outcome across this wider population, e.g. 52 infants from four centres allowed to create a four-variable model of outcome after GM-IVH in (Luyt et al. 2020).

In summary, clinical EEG recordings can index the functional injury after GM-IVH, with higher cortical burst rate reassuring for a positive motor outcome over the first 2 years. This analysis has the potential to provide complementary prognostic information, but also to be used as a cotside non-invasive monitor of cortical health and development, particularly during therapeutic interventions.

## Data Availability

All data produced in the present study are available upon reasonable request to the authors.

## Acknowledgements

This work was supported by Brain Research UK, which had no role in the study design; collection, analysis and interpretation of data; writing of the report; or the decision to submit the article for publication.

We would like to acknowledge the support of the UCL/UCLH Biomedical Research Centre.

## References

Allievi AG, Arichi T, Tusor N, Kimpton J, Arulkumaran S, Counsell SJ, et al. Maturation of Sensori-Motor Functional Responses in the Preterm Brain. Cereb Cortex 2016;26:402–13. https://doi.org/10.1093/cercor/bhv203.

Ancel P-Y, Goffinet F, Kuhn P, Langer B, Matis J, Hernandorena X, et al. Survival and Morbidity of Preterm Children Born at 22 Through 34 Weeks’ Gestation in France in 2011: Results of the EPIPAGE-2 Cohort Study. JAMA Pediatr 2015;169:230–8. https://doi.org/10.1001/jamapediatrics.2014.3351.

Antony JW, Piloto L, Wang M, Pacheco P, Norman KA, Paller KA. Sleep Spindle Refractoriness Segregates Periods of Memory Reactivation. Curr Biol 2018;28:1736–1743.e4. https://doi.org/10.1016/j.cub.2018.04.020.

Arkilo D, Wang S, Thiele EA. Time interval required for return to baseline of the background rhythm on electroencephalogram after recorded electrographic seizures. Epilepsy Res 2013;106:288–91. https://doi.org/10.1016/j.eplepsyres.2013.04.007.

Aso K, Abdab-Barmada M, Scher MS. EEG and the neuropathology in premature neonates with intraventricular hemorrhage. J Clin Neurophysiol 1993;10:304–13.

Aso K, Scher MS, Barmada MA. Neonatal electroencephalography and neuropathology. J Clin Neurophysiol 1989;6:103–23. https://doi.org/10.1097/00004691-198904000-00001.

Bell AH, Greisen G, Pryds O. Comparison of the effects of phenobarbitone and morphine administration on EEG activity in preterm babies. Acta Paediatr 1993;82:35–9. https://doi.org/10.1111/j.1651-2227.1993.tb12511.x.

Benders MJ, Palmu K, Menache C, Borradori-Tolsa C, Lazeyras F, Sizonenko S, et al. Early Brain Activity Relates to Subsequent Brain Growth in Premature Infants. Cereb Cortex 2015;25:3014–24. https://doi.org/10.1093/cercor/bhu097.

Bolisetty S, Dhawan A, Abdel-Latif M, Bajuk B, Stack J, Oei J-L, et al. Intraventricular Hemorrhage and Neurodevelopmental Outcomes in Extreme Preterm Infants. Pediatrics 2014;133:55–62. https://doi.org/10.1542/peds.2013-0372.

Chalak LF, Sikes NC, Mason MJ, Kaiser JR. Low-Voltage aEEG as Predictor of Intracranial Hemorrhage in Preterm Infants. Pediatr Neurol 2011;44:364–9. https://doi.org/10.1016/j.pediatrneurol.2010.11.018.

Cizmeci MN, de Vries LS, Ly LG, van Haastert IC, Groenendaal F, Kelly EN, et al. Periventricular Hemorrhagic Infarction in Very Preterm Infants: Characteristic Sonographic Findings and Association with Neurodevelopmental Outcome at Age 2 Years. J Pediatr 2020;217:79–85.e1. https://doi.org/10.1016/j.jpeds.2019.09.081.

Conde JRC, Hoyos ALR de, Martínez ED, Campo CG, Pérez AM, Hernández Borges AA. Extrauterine life duration and ontogenic EEG parameters in preterm newborns with and without major ultrasound brain lesions. Clin Neurophysiol 2005;116:2796–809. https://doi.org/10.1016/j.clinph.2005.08.020.

Connell J, Vries L de, Oozeer R, Regev R, Dubowitz LMS, Dubowitz V. Predictive Value of Early Continuous Electroencephalogram Monitoring in Ventilated Preterm Infants With Intraventricular Hemorrhage. Pediatrics 1988;82:337–43.

De Wel O, Van Huffel S, Lavanga M, Jansen K, Dereymaeker A, Dudink J, et al. Relationship Between Early Functional and Structural Brain Developments and Brain Injury in Preterm Infants. Cerebellum 2021. https://doi.org/10.1007/s12311-021-01232-z.

Delorme A, Makeig S. EEGLAB: an open source toolbox for analysis of single-trial EEG dynamics including independent component analysis. J Neurosci Methods 2004;134:9–21. https://doi.org/10.1016/j.jneumeth.2003.10.009.

Fabrizi L, Slater R, Worley A, Meek J, Boyd S, Olhede S, et al. A Shift in Sensory Processing that Enables the Developing Human Brain to Discriminate Touch from Pain. Curr Biol 2011;21:1552–8. https://doi.org/10.1016/j.cub.2011.08.010.

Gale C, Statnikov Y, Jawad S, Uthaya SN, Modi N. Neonatal brain injuries in England: population-based incidence derived from routinely recorded clinical data held in the National Neonatal Research Database. Arch Dis Child Fetal Neonatal Ed 2018;103:F301–6. https://doi.org/10.1136/archdischild-2017-313707.

Georgoulas A, Jones L, Laudiano-Dray MP, Meek J, Fabrizi L, Whitehead K. Sleep–wake regulation in preterm and term infants. Sleep 2021;44. https://doi.org/10.1093/sleep/zsaa148.

Golbs A, Nimmervoll B, Sun J-J, Sava IE, Luhmann HJ. Control of Programmed Cell Death by Distinct Electrical Activity Patterns. Cereb Cortex 2011;21:1192–202. https://doi.org/10.1093/cercor/bhq200.

Greisen G, Hellström-Westas L, Lou H, Rosén I, Svenningsen NW. EEG Depression and Germinal Layer Haemorrhage in the Newborn. Acta Pædiatr 1987;76:519–25. https://doi.org/10.1111/j.1651-2227.1987.tb10509.x.

Guzzetta A, Baldini S, Bancale A, Baroncelli L, Ciucci F, Ghirri P, et al. Massage Accelerates Brain Development and the Maturation of Visual Function. J Neurosci 2009;29:6042–51.

Hanganu IL, Staiger JF, Ben-Ari Y, Khazipov R. Cholinergic Modulation of Spindle Bursts in the Neonatal Rat Visual Cortex In Vivo. J Neurosci 2007;27:5694–705.

Hartley C, Berthouze L, Mathieson SR, Boylan GB, Rennie JM, Marlow N, et al. Long-Range Temporal Correlations in the EEG Bursts of Human Preterm Babies. PLOS ONE 2012;7:e31543. https://doi.org/10.1371/journal.pone.0031543.

Hellström-Westas L, Klette H, Thorngren-Jerneck K, Rosén I. Early Prediction of Outcome with aEEG in Preterm Infants with Large Intraventricular Hemorrhages. Neuropediatrics 2001;32:319–24. https://doi.org/10.1055/s-2001-20408.

Hellström-Westas L, Rosén I, Svenningsen NW. Cerebral Function Monitoring During the First Week of Life in Extremely Small Low Birthweight (ESLBW) Infants. Neuropediatrics 1991;22:27–32. https://doi.org/10.1055/s-2008-1071411.

Hollebrandse NL, Spittle AJ, Burnett AC, Anderson PJ, Roberts G, Doyle LW, et al. School-age outcomes following intraventricular haemorrhage in infants born extremely preterm. Arch Dis Child Fetal Neonatal Ed 2021;106:4–8. https://doi.org/10.1136/archdischild-2020-318989.

van den Hoogen A, Teunis CJ, Shellhaas RA, Pillen S, Benders M, Dudink J. How to improve sleep in a neonatal intensive care unit: A systematic review. Early Hum Dev 2017;113:78–86. https://doi.org/10.1016/j.earlhumdev.2017.07.002.

Horst HT, Olffen MV, Remmelts HJ, Vries HD, Bos AF. The prognostic value of amplitude integrated EEG in neonatal sepsis and/or meningitis. Acta Paediatr 2010;99:194–200. https://doi.org/10.1111/j.1651-2227.2009.01567.x.

Iyer KK, Roberts JA, Hellström-Westas L, Wikström S, Pupp IH, Ley D, et al. Cortical burst dynamics predict clinical outcome early in extremely preterm infants. Brain 2015:awv129. https://doi.org/10.1093/brain/awv129.

Kidokoro H, Anderson PJ, Doyle LW, Woodward LJ, Neil JJ, Inder TE. Brain Injury and Altered Brain Growth in Preterm Infants: Predictors and Prognosis. Pediatrics 2014;134:e444–53. https://doi.org/10.1542/peds.2013-2336.

Koskela T, Georgoulas A, Tamuri A, Whitehead K. UCL/NeonatalSleepWake: EEG periodicity analysis 2021a. https://doi.org/10.5281/zenodo.4475695.

Koskela T, Kendall GS, Memon S, Sokolska M, Mabuza T, Huertas-Ceballos A, et al. Prognostic value of neonatal EEG following therapeutic hypothermia in survivors of hypoxic-ischemic encephalopathy. Clin Neurophysiol 2021b;132:2091–100. https://doi.org/10.1016/j.clinph.2021.05.031.

Lamblin M-D, Walls Esquivel E, André M. The electroencephalogram of the full-term newborn: Review of normal features and hypoxic-ischemic encephalopathy patterns. Neurophysiol Clin 2013;43:267–87. https://doi.org/10.1016/j.neucli.2013.07.001.

Lebedeva J, Zakharov A, Ogievetsky E, Minlebaeva A, Kurbanov R, Gerasimova E, et al. Inhibition of Cortical Activity and Apoptosis Caused by Ethanol in Neonatal Rats In Vivo. Cereb Cortex 2017;27:1068–82. https://doi.org/10.1093/cercor/bhv293.

Leijser LM, Miller SP, Wezel-Meijler G van, Brouwer AJ, Traubici J, Haastert IC van, et al. Posthemorrhagic ventricular dilatation in preterm infants: When best to intervene? Neurology 2018;90:e698–706. https://doi.org/10.1212/WNL.0000000000004984.

Leikos S, Tokariev A, Koolen N, Nevalainen P, Vanhatalo S. Cortical responses to tactile stimuli in preterm infants. Eur J Neurosci 2020;51:1059–73. https://doi.org/10.1111/ejn.14613.

Leroy-Terquem E, Vermersch AI, Dean P, Assaf Z, Boddaert N, Lapillonne A, et al. Abnormal Interhemispheric Synchrony in Neonatal Hypoxic-Ischemic Encephalopathy: A Retrospective Pilot Study. NEO 2017;112:359–64. https://doi.org/10.1159/000478964.

Linder N, Haskin O, Levit O, Klinger G, Prince T, Naor N, et al. Risk Factors for Intraventricular Hemorrhage in Very Low Birth Weight Premature Infants: A Retrospective Case-Control Study. Pediatrics 2003;111:e590–5. https://doi.org/10.1542/peds.111.5.e590.

Marcano-Reik AJ, Prasad T, Weiner JA, Blumberg MS. An abrupt developmental shift in callosal modulation of sleep-related spindle bursts coincides with the emergence of excitatory-inhibitory balance and a reduction of somatosensory cortical plasticity. Behav Neurosci 2010;124:600–11. https://doi.org/10.1037/a0020774.

Marret S, Marchand-Martin L, Picaud J-C, Hascoët J-M, Arnaud C, Rozé J-C, et al. Brain Injury in Very Preterm Children and Neurosensory and Cognitive Disabilities during Childhood: The EPIPAGE Cohort Study. PLOS ONE 2013;8:e62683. https://doi.org/10.1371/journal.pone.0062683.

Minlebaev M, Ben-Ari Y, Khazipov R. NMDA Receptors Pattern Early Activity in the Developing Barrel Cortex In Vivo. Cereb Cortex 2009;19:688–96. https://doi.org/10.1093/cercor/bhn115.

Molnár Z, Luhmann HJ, Kanold PO. Transient cortical circuits match spontaneous and sensory-driven activity during development. Science 2020;370. https://doi.org/10.1126/science.abb2153.

Mukerji A, Shah V, Shah PS. Periventricular/Intraventricular Hemorrhage and Neurodevelopmental Outcomes: A Meta-analysis. Pediatrics 2015;136:1132–43. https://doi.org/10.1542/peds.2015-0944.

Olischar M, Klebermass K, Waldhoer T, Pollak A, Weninger M. Background patterns and sleep-wake cycles on amplitude-integrated electroencephalography in preterms younger than 30 weeks gestational age with peri-/intraventricular haemorrhage. Acta Paediatr 2007;96:1743–50. https://doi.org/10.1111/j.1651-2227.2007.00462.x.

Omidvarnia A, Fransson P, Metsäranta M, Vanhatalo S. Functional Bimodality in the Brain Networks of Preterm and Term Human Newborns. Cereb Cortex 2014;24:2657–68. https://doi.org/10.1093/cercor/bht120.

Omidvarnia A, Metsäranta M, Lano A, Vanhatalo S. Structural damage in early preterm brain changes the electric resting state networks. NeuroImage 2015;120:266–73. https://doi.org/10.1016/j.neuroimage.2015.06.091.

Osredkar D, Toet MC, Rooij LGM van, Huffelen AC van, Groenendaal F, Vries LS de. Sleep-Wake Cycling on Amplitude-Integrated Electroencephalography in Term Newborns With Hypoxic-Ischemic Encephalopathy. Pediatrics 2005;115:327–32. https://doi.org/10.1542/peds.2004-0863.

Ouwehand S, Smidt LCA, Dudink J, Benders MJNL, de Vries LS, Groenendaal F, et al. Predictors of Outcomes in Hypoxic-Ischemic Encephalopathy following Hypothermia: A Meta-Analysis. NEO 2020:1–17. https://doi.org/10.1159/000505519.

Patra K, Wilson-Costello D, Taylor HG, Mercuri-Minich N, Hack M. Grades I-II intraventricular hemorrhage in extremely low birth weight infants: Effects on neurodevelopment. J Pediatr 2006;149:169–73. https://doi.org/10.1016/j.jpeds.2006.04.002.

Payne ET, Zhao XY, Frndova H, McBain K, Sharma R, Hutchison JS, et al. Seizure burden is independently associated with short term outcome in critically ill children. Brain 2014;137:1429–38. https://doi.org/10.1093/brain/awu042.

Pisani F, Barilli AL, Sisti L, Bevilacqua G, Seri S. Preterm infants with video-EEG confirmed seizures: Outcome at 30 months of age. Brain Dev 2008;30:20–30. https://doi.org/10.1016/j.braindev.2007.05.003.

Pisani F, Facini C, Bianchi E, Giussani G, Piccolo B, Beghi E. Incidence of neonatal seizures, perinatal risk factors for epilepsy and mortality after neonatal seizures in the province of Parma, Italy. Epilepsia 2018;59:1764–73. https://doi.org/10.1111/epi.14537.

Pressler RM, Cilio MR, Mizrahi EM, Moshé SL, Nunes ML, Plouin P, et al. The ILAE classification of seizures and the epilepsies: Modification for seizures in the neonate. Position paper by the ILAE Task Force on Neonatal Seizures. Epilepsia 2021;62:615–28. https://doi.org/10.1111/epi.16815.

Radvanyi-Bouvet M-F, de Bethmann O, Monset-Couchard M, Fazzi E. Cerebral lesions in early prematurity: EEG prognostic value in the neonatal period. Brain Dev 1987;9:399–405. https://doi.org/10.1016/S0387-7604(87)80113-4.

Ranasinghe S, Or G, Wang EY, Ievins A, McLean MA, Niell CM, et al. Reduced Cortical Activity Impairs Development and Plasticity after Neonatal Hypoxia Ischemia. J Neurosci 2015;35:11946–59. https://doi.org/10.1523/JNEUROSCI.2682-14.2015.

Scher MS, Aso K, Beggarly ME, Hamid MY, Steppe DA, Painter MJ. Electrographic Seizures in Preterm and Full-Term Neonates: Clinical Correlates, Associated Brain Lesions, and Risk for Neurologic Sequelae. Pediatrics 1993;91:128–34.

Tataranno ML, Claessens NHP, Moeskops P, Toet MC, Kersbergen KJ, Buonocore G, et al. Changes in brain morphology and microstructure in relation to early brain activity in extremely preterm infants. Pediatr Res 2018;83:834–42. https://doi.org/10.1038/pr.2017.314.

Tataranno ML, Gui L, Hellström-Westas L, Toet M, Groenendaal F, Claessens NHP, et al. Morphine affects brain activity and volumes in preterms: An observational multi-center study. Early Hum Dev 2020;144:104970. https://doi.org/10.1016/j.earlhumdev.2020.104970.

Thorp JA, Jones PG, Clark RH, Knox E, Peabody JL. Perinatal factors associated with severe intracranial hemorrhage. Am J Obstet Gynecol 2001;185:859–62. https://doi.org/10.1067/mob.2001.117355.

Tokariev A, Palmu K, Lano A, Metsäranta M, Vanhatalo S. Phase synchrony in the early preterm EEG: Development of methods for estimating synchrony in both oscillations and events. NeuroImage 2012;60:1562–73. https://doi.org/10.1016/j.neuroimage.2011.12.080.

Tolner EA, Sheikh A, Yukin AY, Kaila K, Kanold PO. Subplate Neurons Promote Spindle Bursts and Thalamocortical Patterning in the Neonatal Rat Somatosensory Cortex. J Neurosci 2012;32:692–702. https://doi.org/10.1523/JNEUROSCI.1538-11.2012.

Tortora D, Martinetti C, Severino M, Uccella S, Malova M, Parodi A, et al. The effects of mild germinal matrix-intraventricular haemorrhage on the developmental white matter microstructure of preterm neonates: a DTI study. Eur Radiol 2018;28:1157–66. https://doi.org/10.1007/s00330-017-5060-0.

Vanhatalo S, Palva JM, Andersson S, Rivera C, Voipio J, Kaila K. Slow endogenous activity transients and developmental expression of K+–Cl− cotransporter 2 in the immature human cortex. Eur J Neurosci 2005;22:2799–804. https://doi.org/10.1111/j.1460-9568.2005.04459.x.

Vasileiadis GT, Gelman N, Han VKM, Williams L-A, Mann R, Bureau Y, et al. Uncomplicated Intraventricular Hemorrhage Is Followed by Reduced Cortical Volume at Near-Term Age. Pediatrics 2004;114:e367–72. https://doi.org/10.1542/peds.2004-0500.

Vries L de, Liem KD, Dijk K van, Smit BJ, Sie L, Rademaker KJ, et al. Early versus late treatment of posthaemorrhagic ventricular dilatation: results of a retrospective study from five neonatal intensive care units in The Netherlands. Acta Paediatr 2002;91:212–7. https://doi.org/10.1111/j.1651-2227.2002.tb01697.x.

de Vries LS. Hemorrhagic Lesions of the Central Nervous System. Fetal and Neonatal Brain Injury. Fifth, Cambridge University Press; 2018, p. 247–57.

Whitehead K, Jones L, Laudiano-Dray MP, Meek J, Fabrizi L. Altered cortical processing of somatosensory input in pre-term infants who had high-grade germinal matrix-intraventricular haemorrhage. NeuroImage Clin 2020;25:102095. https://doi.org/10.1016/j.nicl.2019.102095.

Whitehead K, Papadelis C, Laudiano-Dray MP, Meek J, Fabrizi L. The Emergence of Hierarchical Somatosensory Processing in Late Prematurity. Cereb Cortex 2019;29:2245–60. https://doi.org/10.1093/cercor/bhz030.

Whitehead K, Pressler R, Fabrizi L. Characteristics and clinical significance of delta brushes in the EEG of premature infants. Clin Neurophysiol Pract 2016;2:12–8. https://doi.org/10.1016/j.cnp.2016.11.002.

Whitehead K, Rupawala M, Laudiano-Dray MP, Meek J, Olhede S, Fabrizi L. Spontaneous activation of cortical somatosensory networks depresses their excitability in preterm human neonates 2022:2022.12.08.519675. https://doi.org/10.1101/2022.12.08.519675.

