## Supplementary Information for "Clinical value of cortical bursting in preterm infants with intraventricular haemorrhage"

**Supplementary Methods**

EEG data were either recorded from 0.053-500Hz with a FCz reference using the NicoletOne recording system and digitized with a sampling rate of 250 or 500Hz, or from 0.08-450Hz with a CPz reference using the Micromed recording system and digitized with a sampling rate of 256Hz. Therefore, to make data consistent, they were filtered with a high-passband edge of 0.1Hz, down-sampled to 250Hz, and re-referenced to Cz using EEGLAB (Swartz Center for Computational Neuroscience) (in recordings without Cz, this channel was estimated with spherical interpolation as implemented in EEGLAB).

**Supplementary Figures**

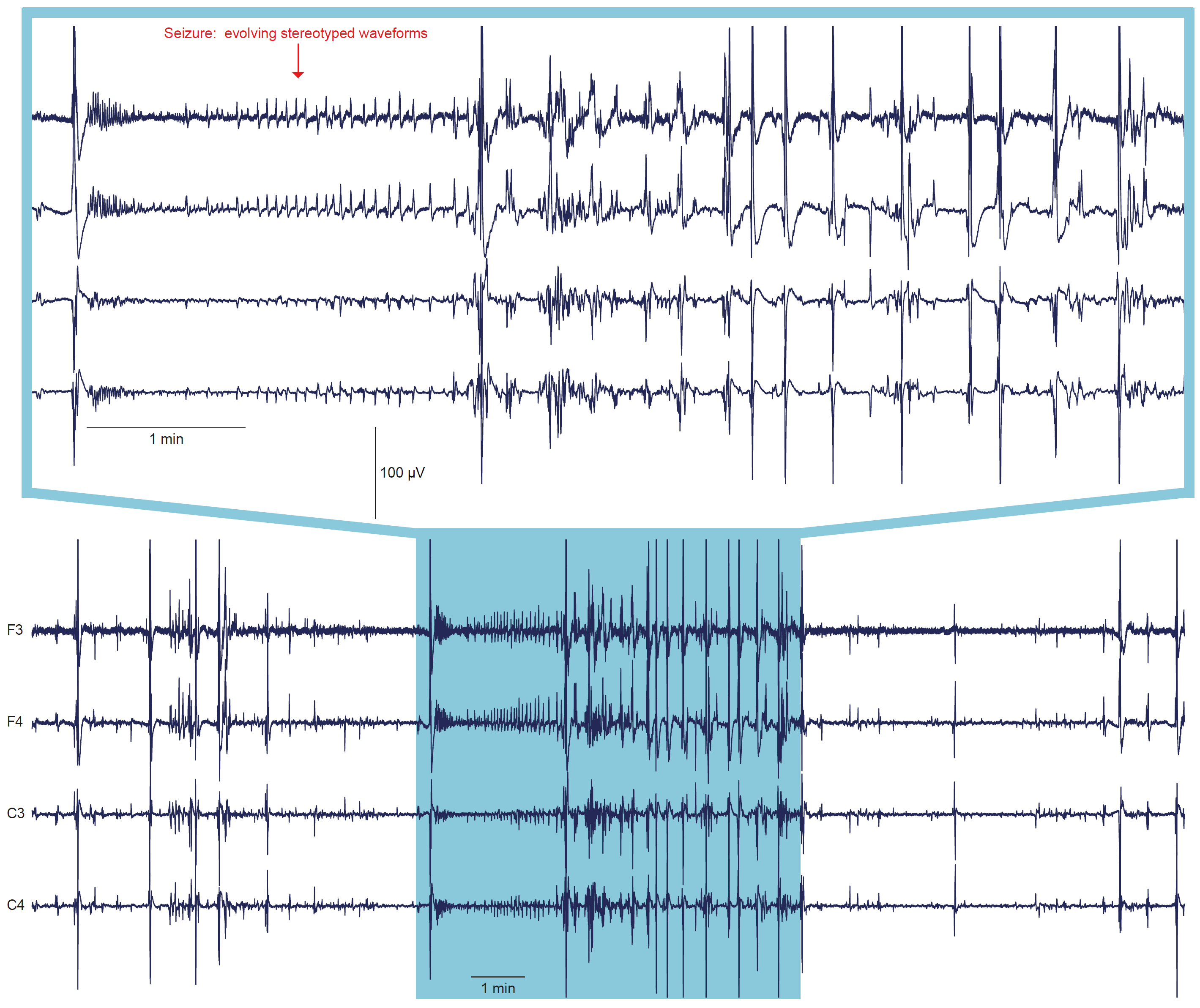

**Supplementary Fig. 1: Seizure in infant #9 at postnatal day 4 and corrected gestational age 25+4 weeks.** No display filters applied.

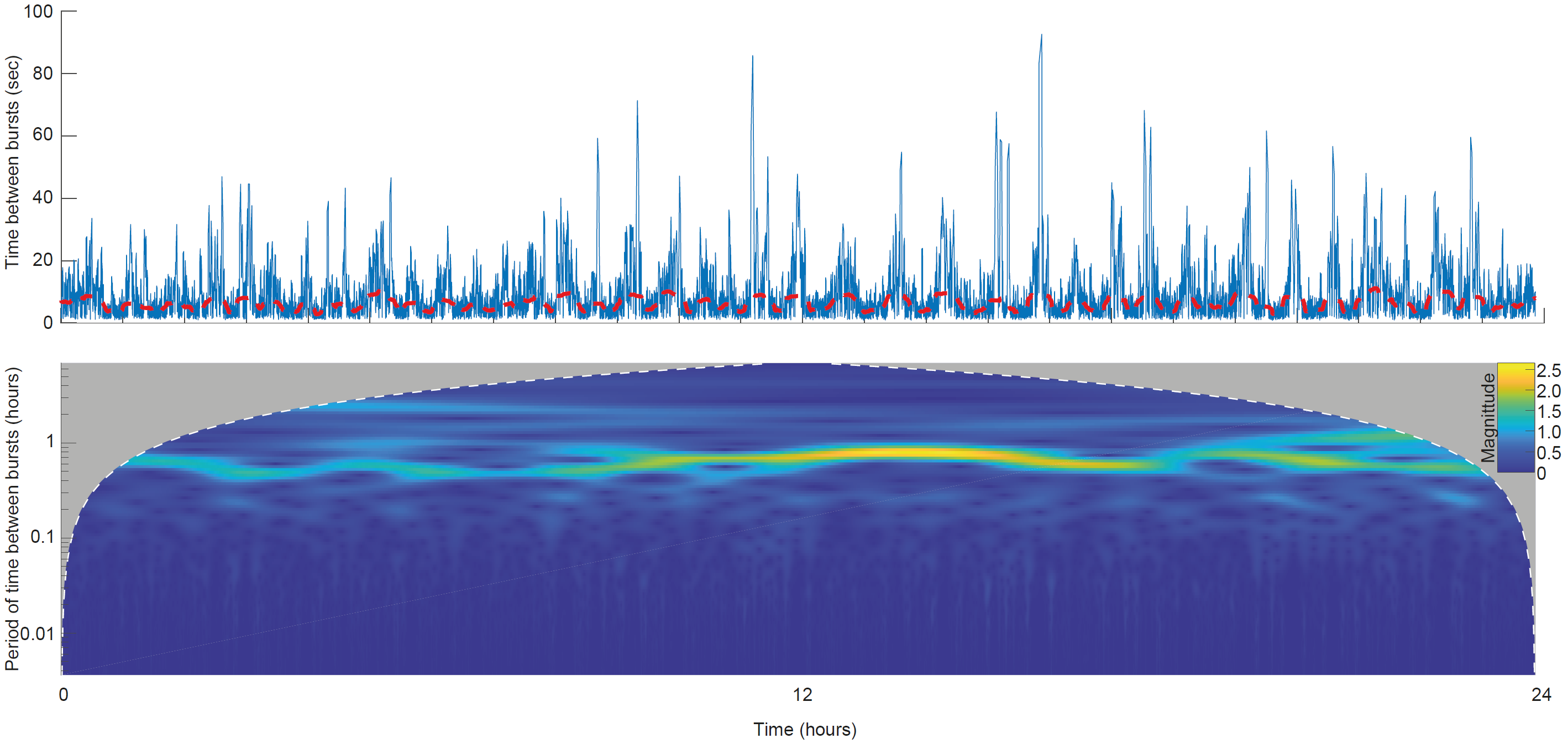

**Supplementary Fig. 2: Burst occurrence rate could be periodic in very preterm infants.** Upper panel: time series of time since last burst at a representative (central) channel for infant #798 at postnatal day 3 and corrected gestational age 31+6 weeks+days. Lower panel: continuous wavelet transform of time since last burst (the data plotted in the upper panel). Note the high magnitude periodicity with an approximately 1 hour cycle sustained across 24 hours, denoted by the green/yellow horizontal ‘streak’. These data support the presence of sleep cycling: the periods of higher burst rate/shorter time between bursts are consistent with cycles of active sleep (Watanabe and Iwase, 1972; Whitehead et al., 2016).

**References**

Watanabe, K., Iwase, K., 1972. Spindle-like Fast Rhythms in the EEGs of Low-birthweight Infants. Dev Med Child Neurol 14, 373–381. https://doi.org/10.1111/j.1469-8749.1972.tb02603.x

Whitehead, K., Pressler, R., Fabrizi, L., 2016. Characteristics and clinical significance of delta brushes in the EEG of premature infants. Clin Neurophysiol Pract 2, 12–18. https://doi.org/10.1016/j.cnp.2016.11.002
